# Pre-migratory, Migratory and Post-migratory Factors Associated with Risky Sexual Behaviour among Women Informal Cross-Border Traders at the COMESA Market, Lusaka, Zambia

**DOI:** 10.64898/2026.07.22.26358726

**Authors:** Simson Mwale, Musonda Lemba, Million Phiri

## Abstract

Women engaged in informal cross-border trade in Southern Africa experience frequent mobility, economic pressure, and exposure to new social environments that may increase vulnerability to risky sexual behaviour and HIV. Limited evidence exists on how pre-migratory, migratory, and post-migratory factors interact to shape sexual risk among this population. This study examined the prevalence and determinants of risky sexual behaviour among women informal cross-border traders at COMESA Market in Lusaka, Zambia. A cross-sectional survey was conducted with 499 women informal cross-border traders. A composite risk score was used to classify respondents as “low risk” or “high risk” for sexual behaviour. Descriptive statistics, chi-square tests, and multivariate binary logistic regression were used to investigate associations between selected covariates and risky sexual behaviour, guided by migration and vulnerability theory. The majority (67.7%) of respondents were classified as high-risk. In multivariate analysis, women who used personal savings for start-up capital were 73% less likely to be high-risk compared to those relying on nuclear family capital (aOR = 0.27, 95% CI: 0.10-0.73). Women with 5-9 years of trading experience also had lower odds (aOR = 0.21, 95% CI: 0.29-0.96). Conversely, using Katima Mulilo border increased the odds 15-fold (aOR = 14.59, 95% CI: 1.26-169.50). Not feeling disconnected while away (aOR = 3.39, 95% CI: 1.03-11.11) and sharing accommodation with one roommate (aOR = 4.42, 95% CI: 1.10-17.84) were also significant predictors of high-risk behaviour. This study demonstrates that risk among women informal cross-border traders is produced across the migration continuum by economic dependence, specific transit routes, and destination living conditions. Interventions should therefore prioritise financial independence, targeted health and protection services at high-risk borders, and safe accommodation with peer-led HIV prevention to reduce vulnerability among mobile women traders.

## Introduction

International migration and mobility are increasingly recognised as key social determinants of health, particularly for women engaged in informal economic activities across borders [1]. In the Common Market for Eastern and Southern Africa (COMESA) region, informal cross-border trade is a critical livelihood strategy, with women constituting the majority of traders who move frequently between countries to buy and sell goods [2,3,18]. In Zambia, the COMESA Market in Lusaka serves as a major hub where women traders source and sell merchandise from Tanzania, Democratic Republic of the Congo, Malawi, South Africa, and other neighbouring countries. While informal cross-border trade provides essential income for households and contributes to regional trade [4,5], it requires repeated travel, extended periods away from home, and engagement in unfamiliar social and economic environments that may heighten health vulnerabilities [6,7].

Evidence from migration and health literature shows that mobility can increase exposure to sexual risk through the disruption of social networks, economic pressure, and contact with new sexual partners [1,8,14,15]. For women informal cross-border traders, these risks are compounded by low and irregular incomes, dependence on family or informal sources for start-up capital, and the need to stay in low-cost, shared accommodation during trade trips [9,10]. Yet, despite the large number of women involved in informal cross-border trade, there remains limited empirical data on how mobility-related factors shape sexual health outcomes for this population in Zambia. Most existing studies have focused on long-term migrants or sex workers, leaving a gap in understanding the specific risks faced by women whose movement is driven by trade [11].

This study is guided by migration and vulnerability theory, which argues that health risks among mobile populations are not inherent but are produced across three interrelated stages of migration [1,8,19]. Pre-migratory factors, such as economic resources and household responsibilities at origin, determine the level of autonomy and pressure to trade. Migratory factors, including travel frequency, duration away, and specific border crossing points, shape exposure during transit [9.13]. Post-migratory factors, such as type of accommodation, social integration, and feelings of isolation at destination, influence health behaviours in the trading environment [12,13]. Within this framework, vulnerability to risky sexual behaviour emerges from the interaction between individual characteristics and the structural, social, and economic contexts encountered before, during, and after migration. For women informal cross-border traders, financial dependence may limit bargaining power, certain border routes may present higher risk environments, and shared living conditions may weaken social controls and reinforce peer norms that normalise risk [7,9].

At COMESA Market, thousands of women undertake repeated trading trips, often supporting dependents and staying in lodges or hostels with other traders [2,19]. National data also show persistent HIV burden in Zambia, with mobility identified as a key driver [14,15]. Preliminary observations suggest high economic pressure and frequent mobility, but the extent to which pre-migratory economic factors, migratory patterns, and post-migratory living conditions contribute to risky sexual behaviour has not been documented. Without this evidence, HIV prevention programmes risk remaining generic and failing to address the structural drivers of vulnerability that operate across the migration continuum.

Therefore, this study aimed to determine the prevalence and factors associated with risky sexual behaviour among women informal cross-border traders at COMESA Market in Lusaka. More specifically, it examined how selected pre-migratory, migratory, and post-migratory characteristics relate to high-risk sexual behaviour. The study sought to provide evidence to inform targeted HIV prevention and protection interventions that promote financial autonomy, safer mobility, and health protection for a highly mobile and economically important, yet understudied population, by applying a migration and vulnerability lens.

## Methods

### Study Design and Setting

This cross-sectional study was conducted among women informal cross-border traders at COMESA Market in Lusaka, Zambia. The COMESA Market was established in 1998 by the Cross-Border Traders Association (CBTA) and serves as one of the largest hubs for informal cross-border trade in the region [2]. By 2015, the market had a membership of approximately 42,350 cross-border traders, of whom 70% were women [16]. At the time of the survey, the market comprised 1,299 shops and containers distributed across 14 sections and zones, with 49% (n = 632) owned or rented by women [17]. The market setting was selected because it concentrates a large population of women engaged in informal cross-border trade, whose mobility and socio-economic conditions may influence sexual health outcomes [6,7].

### Study Population and Sampling

The study population consisted of women aged 18 years and above who were members of the CBTA, actively engaged in cross-border trading, and owned or rented a shop or container at the COMESA Market. Women who were not present at the market during the data collection period were excluded. A three-stage multi-stage probability sampling technique was employed to select participants. In the first stage, women traders were identified within specific business sections of the market. In the second stage, stratified sampling was used to select representative trading units from each section and zone. In the third stage, simple random sampling was applied to select individual respondents from the sampled trading units. This approach yielded a final sample of 499 women, representing the population of women informal cross-border traders at the market. The study achieved a response rate of 99.8%. The sampling strategy was designed to ensure representation across the different sections of the market and to focus on women whose demographic and migration characteristics could influence levels of risky sexual behaviour [6].

### Data Collection

Data were collected in January 2019 using structured questionnaires administered by trained female research assistants. The questionnaire captured information on socio-demographic characteristics including marital status, age, education level, literacy, religious affiliation, and place of residence. It also collected self-reported data on sexual behaviours and detailed information on migration experiences. The migration component was organised to capture pre-migratory, migratory, and post-migratory factors relevant to cross-border trade, in order to examine how experiences at the point of departure, during transit, and at the destination relate to sexual risk.

### Study Variables

#### Outcome Variable

The dependent variable for this study was risky sexual behaviour (Table 1). This was measured using a composite index variable, “RISK_LEVEL”, constructed from 12 survey items assessing sexual risk practices: having more than one sexual partner, engages in voluntary sex, had sex with multiple partners in the past 12 months, was not forced to have sex, was paid to have sex, engages in dry sex, did not use a condom, did not suggest the use of a condom, did not use a condom for various reasons, did not use a condom much during the last 12 months, has heard of STIs, understands and has no knowledge of STIs [14,15]. Each item was coded 1 if it indicated a high-risk behaviour and 0 otherwise. A total score was calculated for each respondent, ranging from 0 (risk-free) to 11 (very high-risk). For analytical purposes, the index was recoded into a binary outcome variable, “RISK_STATUS”, where scores of 0–5 were classified as low-risk and scores of 6–11 as high-risk. Of the 499 respondents, 161 (32.3%) were classified as low-risk and 338 (67.7%) as high-risk, with the highest concentration of respondents scoring 7 (28.3%).

**Table 1.** Recoding of the dependent variable.

| Variable measurement | Survey Results, 2019 (N= 499) |  |
| --- | --- | --- |
|  | Number | Percent |
| **RISK_LEVEL** |  |  |
| 0 | 10 | 2 |
| 1 | 6 | 1.2 |
| 2 | 17 | 3.4 |
| 3 | 29 | 5.8 |
| 4 | 42 | 8.4 |
| 5 | 57 | 11.4 |
| 6 | 86 | 17.2 |
| 7 | 141 | 28.3 |
| 8 | 54 | 10.8 |
| 9 | 33 | 6.6 |
| 10 | 21 | 4.2 |
| 11 | 3 | 0.6 |
| **RISK_STATUS** |  |  |
| Low risk | 161 | 32.3 |
| High risk | 338 | 67.7 |

#### Independent Variables

Independent variables were organised around migration stages [1,11] and included 18 variables in total. Pre-migratory characteristics [3,4] comprised eight variables: person who encouraged entry into trade (nuclear family, extended family, friends, self-discovery), source of start-up capital (family, friends, salary, bank credit, entrepreneurial/small business), year of cross-border trading debut (<1999, 2000–2004, 2005–2009, 2010–2014, 2015+), age at debut (≤24 years, 25–34 years, ≥35 years), number of years as a trader (<5 years, 5–9 years, 10+ years), having dependents (yes/no), number of dependents (≤5, 6+), and whether a partner accompanies the trader on trips (yes/no). Migratory experiences [9,13] included four variables: border crossing point used (Nakonde, Kasumbalesa, Chirundu, Livingstone, Kazungula, Katima Mulilo, Chipata, Kapiri Mposhi), number of days spent out of the country in the last 12 months (none, ≤5, 6–10, 11+), average number of trips out in the last 12 months (none, 1–5, 6+), and whether sexual favours were requested by public officials during transit (yes/no). Post-migratory experiences [7,10] included six variables: feelings when out of the country (lonely, isolated, stressed, nothing/I am used), feeling disconnected (yes/no), being accompanied by a partner at destination (yes/no), type of accommodation (hostel, hotel, lodge, guest house), number of people sharing a room (none, 1 roommate, 2+ roommates), and current economic situation (financially stable, stable, financially strained).

### Statistical Analysis

Data analysis was conducted at three levels: univariate, bivariate, and multivariate using STATA Version 13. Univariate analysis used descriptive statistics to summarise respondents’ migration experiences, and distribution of risk levels. Bivariate analysis employed chi-square tests to examine associations between each independent variable and risky sexual behaviour. Multivariate analysis used binary logistic regression to identify significant predictors of high-risk sexual behaviour while controlling for potential confounders, pre-migratory characteristics. Adjusted odds ratios (aORs) with 95% confidence intervals (CIs) were reported to indicate the strength and direction of associations. Statistical significance was set at p < 0.05.

### Ethical Considerations

Ethical approval was obtained the University of Zambia’s School of Humanities and Social Sciences Research Ethics Committee (HSSREC: 2018-June-005, No. 153760602331). Respondents’ received an information sheet outlining the study’s purpose and procedures. Written consent was obtained from each respondent before data collection. Questionnaires were conducted in a secure location to maintain confidentiality.

## Results

The pre-migratory characteristics of the 499 women informal cross-border traders are presented in Table 2. With regard to who initiated them into cross-border trade, the largest proportion reported being encouraged by friends (n = 168, 36.21%), followed by self-discovery (n = 153, 32.97%). Nuclear family members accounted for 23.71% (n = 110) of initiators, while extended family members represented the smallest group at 7.22% (n = 33). In terms of source of start-up capital, nearly half of the respondents (n = 221, 46.92%) indicated that their nuclear family provided the capital. Personal savings was the second most common source at 23.57% (n = 111), followed by support from friends at 13.80% (n = 65). Bank credit was accessed by 8.70% (n = 41) of women, and 7.01% (n = 33) received capital from extended family members.

**Table 2.** Pre-migratory characteristics of respondents.

| Background characteristic | Survey Results, 2019 (N= 499) |  |
| --- | --- | --- |
|  | Number | Percent |
| **Initiator of cross-border trade** |  |  |
| Nuclear family member | 110 | 23.71 |
| Extended family member | 33 | 7.22 |
| Friends | 168 | 36.21 |
| Self-discovery | 153 | 32.97 |
| **Source of capital** |  |  |
| Nuclear family | 221 | 46.92 |
| Extended family | 33 | 7.01 |
| Friends | 65 | 13.80 |
| Bank credit | 41 | 8.70 |
| Personal savings | 111 | 23.57 |
| **Year of debut** |  |  |
| 1980-1999 | 25 | 5.24 |
| 2000-2004 | 40 | 8.39 |
| 2005-2009 | 95 | 19.92 |
| 2010-2014 | 170 | 35.64 |
| 2015-2019 | 147 | 30.82 |
| **Age at year of cross-border trade** |  |  |
| <= 24 years | 104 | 22.03 |
| 25-34 years | 228 | 48.31 |
| 34+ years | 140 | 29.66 |
| **Period as cross-border trader** |  |  |
| <= 5 years | 171 | 35.70 |
| 5-9 years | 167 | 34.86 |
| 10+ years | 141 | 29.44 |
| **Dependents** |  |  |
| Yes | 371 | 78.60 |
| No | 101 | 21.40 |
| **Number of dependents** |  |  |
| 1-5 dependents | 233 | 63.84 |
| 6+ dependents | 132 | 36.16 |
| **Partner accompaniment** |  |  |
| Yes | 22 | 5.16 |
| No | 404 | 94.84 |

The majority of respondents began cross-border trading in the last two decades. Specifically, 35.64% (n = 170) started between 2010 and 2014, and 30.82% (n = 147) started between 2015 and 2019. Another 19.92% (n = 95) began between 2005 and 2009, while 8.39% (n = 40) started between 2000 and 2004, and 5.24% (n = 25) had been trading since 1980-1999. Regarding age at debut, almost half of the women (n = 228, 48.31%) started trading between 25 and 34 years, classified as young adulthood. Those who started at 24 years and below comprised 22.03% (n = 104), while 29.66% (n = 140) began at age 35 and above. In terms of trading experience, the sample was relatively evenly distributed: 35.70% (n = 171) had been trading for 5 years or less, 34.86% (n = 167) for 5-9 years, and 29.44% (n = 141) for 10 years or more. The data further show that a large majority of respondents had dependents. Overall, 78.60% (n = 371) reported having dependents, while 21.40% (n = 101) did not. Among those with dependents, 63.84% (n = 233) had between 1 and 5 dependents, and 36.16% (n = 132) had 6 or more dependents. Finally, partner accompaniment during trading trips was rare. Only 5.16% (n = 22) of women reported being accompanied by a partner, while 94.84% (n = 404) travelled alone.

In summary, most women entered cross-border trade through peer networks or independently, relied primarily on nuclear family or personal savings for capital, began trading in the last 10-15 years during young adulthood, and carried significant household responsibilities with limited spousal support during travel.

The migratory and post-migratory characteristics of the 499 women informal cross-border traders are presented in Table 3. In terms of border crossings, Nakonde was the most frequently used border point, reported by 29.03% (n = 137) of respondents. This was followed by Kasumbalesa at 16.31% (n = 77) and Livingstone at 13.98% (n = 66). Kazungula was used by 11.86% (n = 56), while Chirundu and Chipata were each used by 8.90% (n = 42). Katima Mulilo accounted for 7.63% (n = 36) and Kapiri Mposhi was the least used at 3.39% (n = 16).

**Table 3.** Migratory and post-migratory characteristics of respondents.

| Background characteristic | Survey Results, 2019 (N= 499) |  |
| --- | --- | --- |
|  | Number | Percent |
| **Border often used** |  |  |
| Nakonde | 137 | 29.03 |
| Kasumbalesa | 77 | 16.31 |
| Chirundu | 42 | 8.90 |
| Livingstone | 66 | 13.98 |
| Kazungula | 56 | 11.86 |
| Katima Mulilo | 36 | 7.63 |
| Chipata | 42 | 8.90 |
| Kapiri Mposhi | 16 | 3.39 |
| **Days spent away (last 12 months)** |  |  |
| None | 39 | 7.88 |
| <= 4 days | 152 | 30.71 |
| 5-9 days | 249 | 50.30 |
| 10+ days | 55 | 11.11 |
| **Average times out** |  |  |
| Not gone out | 46 | 9.62 |
| 1-5 times | 332 | 69.46 |
| 6+ times | 100 | 20.92 |
| **Sexual favours (public official)** |  |  |
| Yes | 81 | 17.23 |
| No | 389 | 82.77 |
| **Feeling being away from loved ones** |  |  |
| Feel lonely | 247 | 55.26 |
| Feel isolated | 63 | 14.09 |
| Feel stressed | 92 | 20.58 |
| Feel nothing | 45 | 10.07 |
| **Feel disconnected being away** |  |  |
| Yes | 241 | 51.39 |
| No | 228 | 48.61 |
| **Type of accommodation** |  |  |
| A bed in hostel | 96 | 20.65 |
| Single room in hotel | 69 | 14.84 |
| Single room at lodge | 300 | 64.52 |
| **Room sharing** |  |  |
| None | 97 | 23.43 |
| 1 roommate | 93 | 22.46 |
| 2+ roommates | 224 | 54.11 |
| **Current economic condition** |  |  |
| Financially stable | 205 | 43.52 |
| Stable | 211 | 44.80 |
| Financially unstable | 55 | 11.68 |

Mobility patterns in the 12 months before the survey indicate frequent travel. Half of the respondents (50.30%, n = 249) reported spending 5-9 days out of the country, while 30.71% (n = 152) spent 4 days or less. Only 11.11% (n = 55) spent 10 or more days away, and 7.88% (n = 39) reported no travel in the last year. Regarding frequency of travel, 69.46% (n = 332) made 1-5 trips out of the country, 20.92% (n = 100) made 6 or more trips, and 9.62% (n = 46) did not travel out.

During transit, 17.23% (n = 81) of women reported being asked for sexual favours by public officials, while the majority, 82.77% (n = 389), reported no such requests. Emotional experiences while away from home were also common. More than half of the respondents (55.26%, n = 247) reported feeling lonely when away from loved ones. A further 20.58% (n = 92) felt stressed, 14.09% (n = 63) felt isolated, and 10.07% (n = 45) reported feeling nothing as they were used to being away. Correspondingly, 51.39% (n = 241) reported feeling disconnected while away, compared to 48.61% (n = 228) who did not.

Accommodation patterns at the destination showed that the majority stayed in single rooms at lodges (64.52%, n = 300). Another 20.65% (n = 96) stayed in a bed in a hostel, and 14.84% (n = 69) stayed in a single room in a hotel. Room sharing was common, with 54.11% (n = 224) sharing with 2 or more roommates, 22.46% (n = 93) sharing with 1 roommate, and 23.43% (n = 97) having no roommate. Finally, with respect to current economic condition, 44.80% (n = 211) described themselves as “stable”, 43.52% (n = 205) as “financially stable”, and 11.68% (n = 55) as “financially unstable”.

In summary, respondents demonstrated high levels of mobility, primarily through Nakonde, with most trips lasting less than 10 days and occurring 1-5 times per year. A notable minority reported solicitation for sexual favours, and over half experienced feelings of loneliness and disconnection. Most stayed in lodge accommodation and shared rooms, while the majority rated their economic condition as either stable or financially stable.

Table 4 shows the distribution of risky sexual behaviour by selected pre-migratory, migratory and post-migratory characteristics.

**Table 4.**
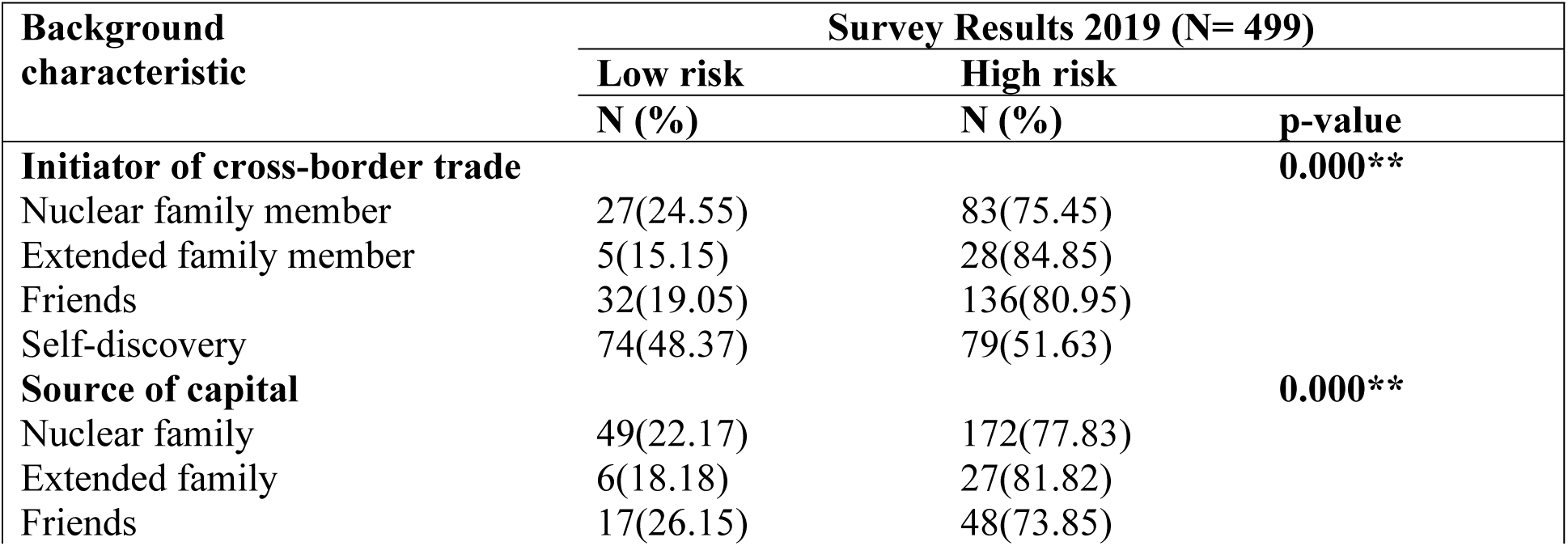
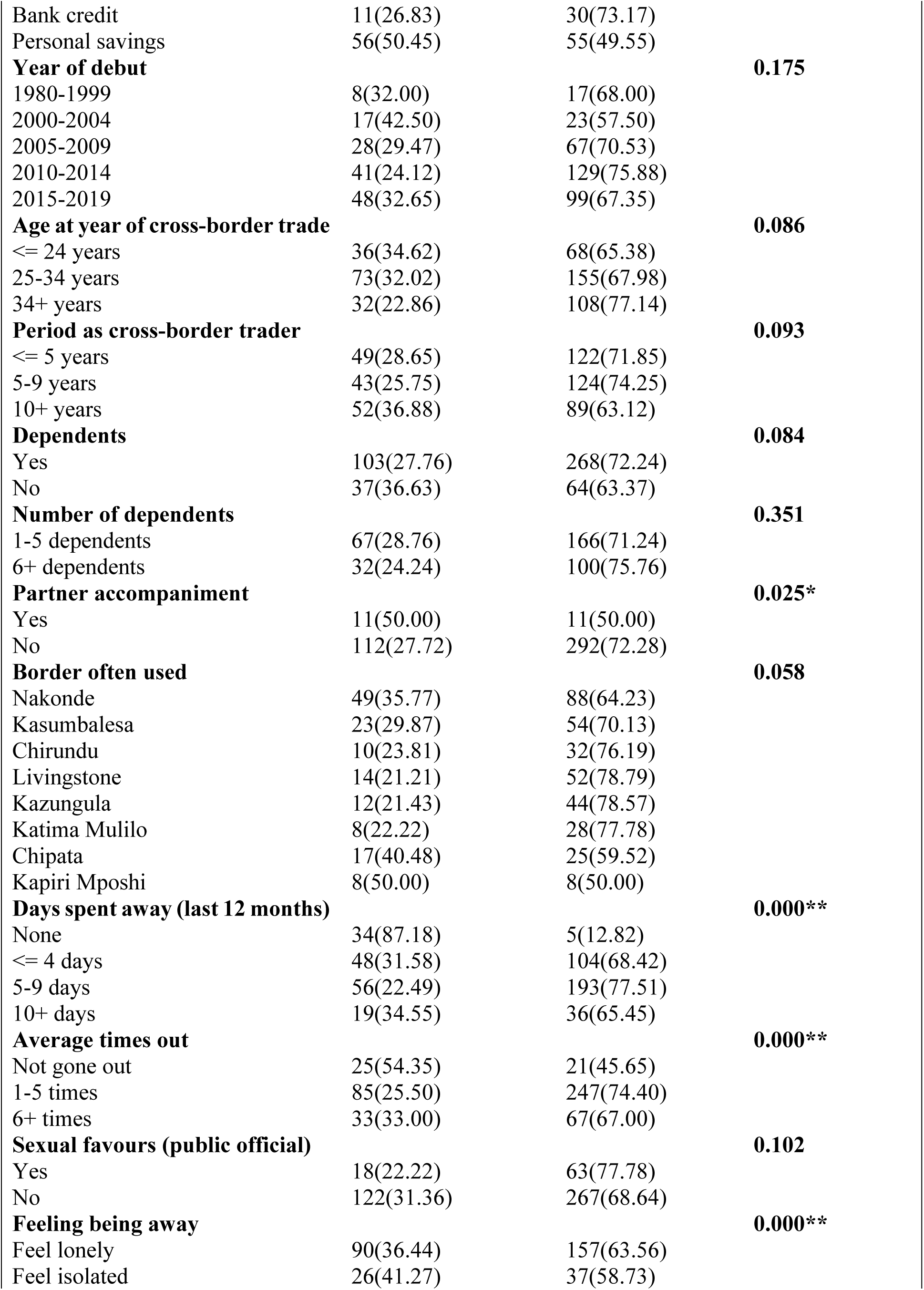
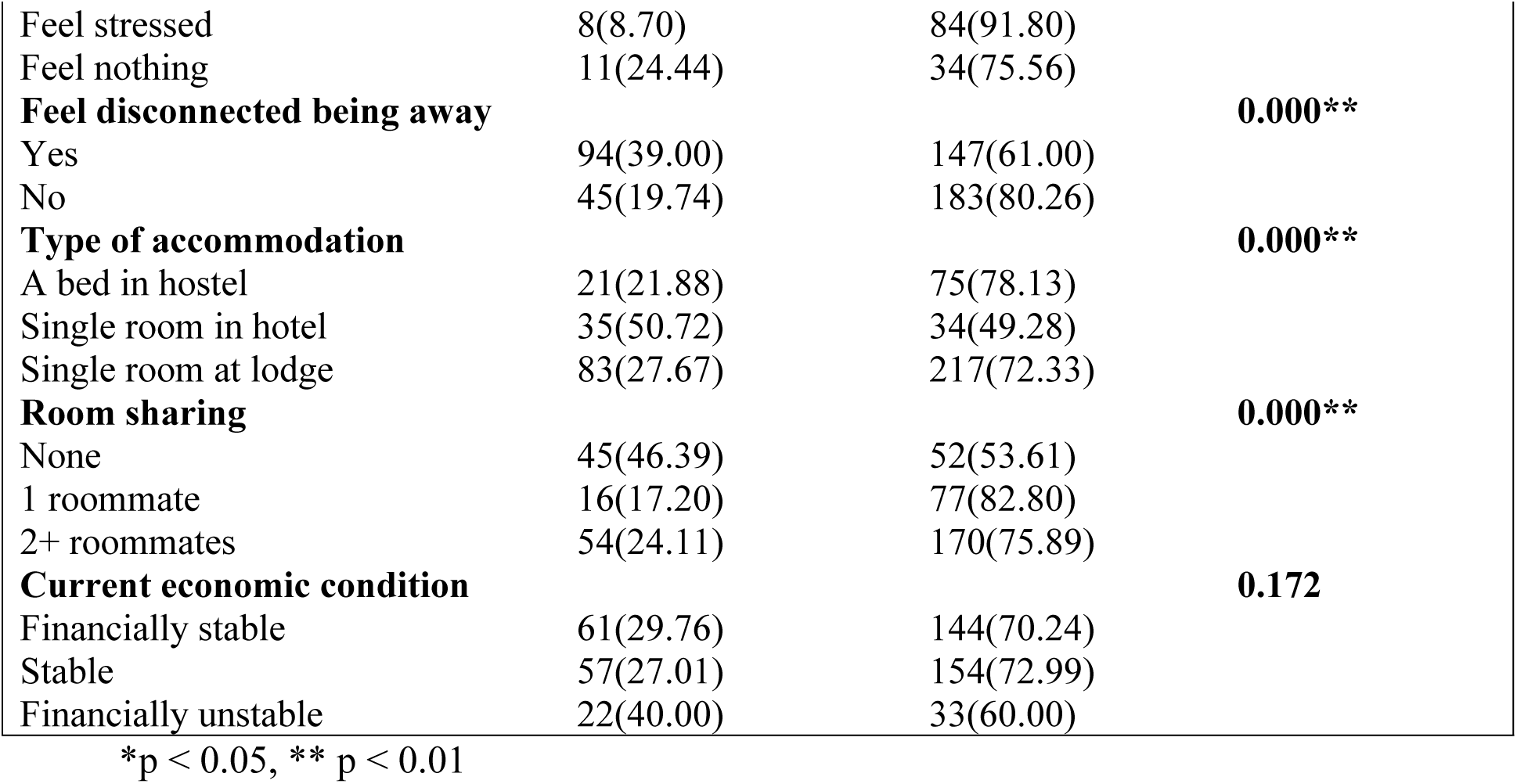
Percent distribution of risky sexual behaviour by selected pre-migratory, migratory and post-migratory characteristics.

With regard to pre-migratory characteristics, risky sexual behaviour varied significantly by who initiated the respondent into cross-border trade (p < 0.01). The highest proportion of high-risk behaviour was reported among those initiated by extended family members (84.85%, n = 28) and friends (80.95%, n = 136), compared to 75.45% (n = 83) among those initiated by nuclear family members and 51.63% (n = 79) among those who entered through self-discovery. Source of start-up capital was also significantly associated with risk status (p < 0.01). Women who received capital from extended family (81.82%, n = 27) and nuclear family (77.83%, n = 172) had the highest levels of high-risk behaviour. In contrast, those who used personal savings had the lowest proportion at 49.55% (n = 55). Partner accompaniment during trips was significantly associated with risk (p < 0.05). Among women who travelled with a partner, 50.00% (n = 11) were classified as high-risk compared to 72.28% (n = 292) among those who travelled alone. Year of debut (p = 0.175), age at debut (p = 0.086), period as trader (p = 0.093), having dependents (p = 0.084), and number of dependents (p = 0.351) were not statistically significantly associated with risky sexual behaviour.

In terms of migratory characteristics, both days spent away in the last 12 months and average number of trips out were strongly associated with risk (p < 0.01). Women who spent 5-9 days away had the highest proportion of high-risk behaviour (77.51%, n = 193), while those who did not travel at all had the lowest (12.82%, n = 5). Similarly, 74.40% (n = 247) of those who travelled 1-5 times and 67.00% (n = 67) of those who travelled 6+ times were high-risk, compared to 45.65% (n = 21) of those who had not gone out. Border point used was not statistically significant (p = 0.058), though high-risk behaviour was highest among users of Kazungula (78.57%, n = 44) and Livingstone (78.79%, n = 52) and lowest among users of Kapiri Mposhi and Chipata. Reports of being asked for sexual favours by public officials were not significantly associated with risk status (p = 0.102), though 77.78% (n = 63) of those who reported such requests were in the high-risk group.

Finally, several post-migratory factors were significantly associated with risky sexual behaviour. Feelings while being away from home were strongly associated with risk (p < 0.01). The highest proportion of high-risk behaviour was among those who felt stressed (91.80%, n = 84), followed by those who felt nothing (75.56%, n = 34) and those who felt lonely (63.56%, n = 157). Feeling disconnected while away was also significant (p < 0.01): 80.26% (n = 183) of women who did not feel disconnected were high-risk compared to 61.00% (n = 147) of those who did. Type of accommodation (p < 0.01) and room sharing (p < 0.01) were also associated with risk. Women staying in a bed in a hostel (78.13%, n = 75) and in a single room at a lodge (72.33%, n = 217) had higher levels of high-risk behaviour than those in a single hotel room (49.28%, n = 34). Regarding room sharing, 82.80% (n = 77) of those sharing with 1 roommate and 75.89% (n = 170) of those sharing with 2+ roommates were high-risk, compared to 53.61% (n = 52) of those with no roommate. Current economic condition was not significantly associated with risk status (p = 0.172).

Overall, the bivariate results suggested that initiation pathways, source of capital, mobility patterns, emotional experiences while away, accommodation type and room sharing were key factors associated with risky sexual behaviour among women informal cross-border traders at COMESA Market.

Table 5 presents results from the binary logistic regression model examining the association between pre-migratory, migratory and post-migratory covariates and the likelihood of engaging in high-risk sexual behaviour.

**Table 5.**
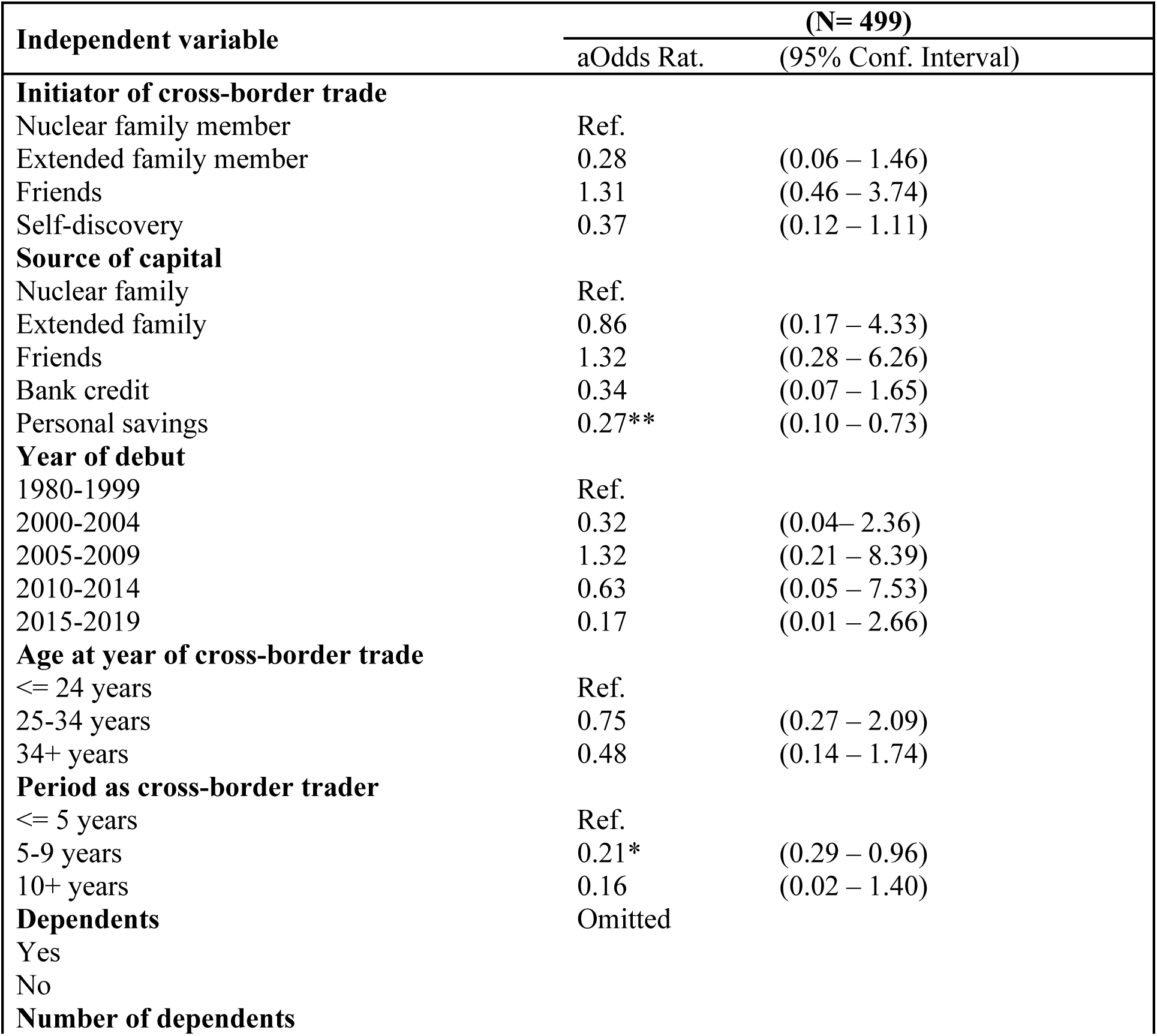
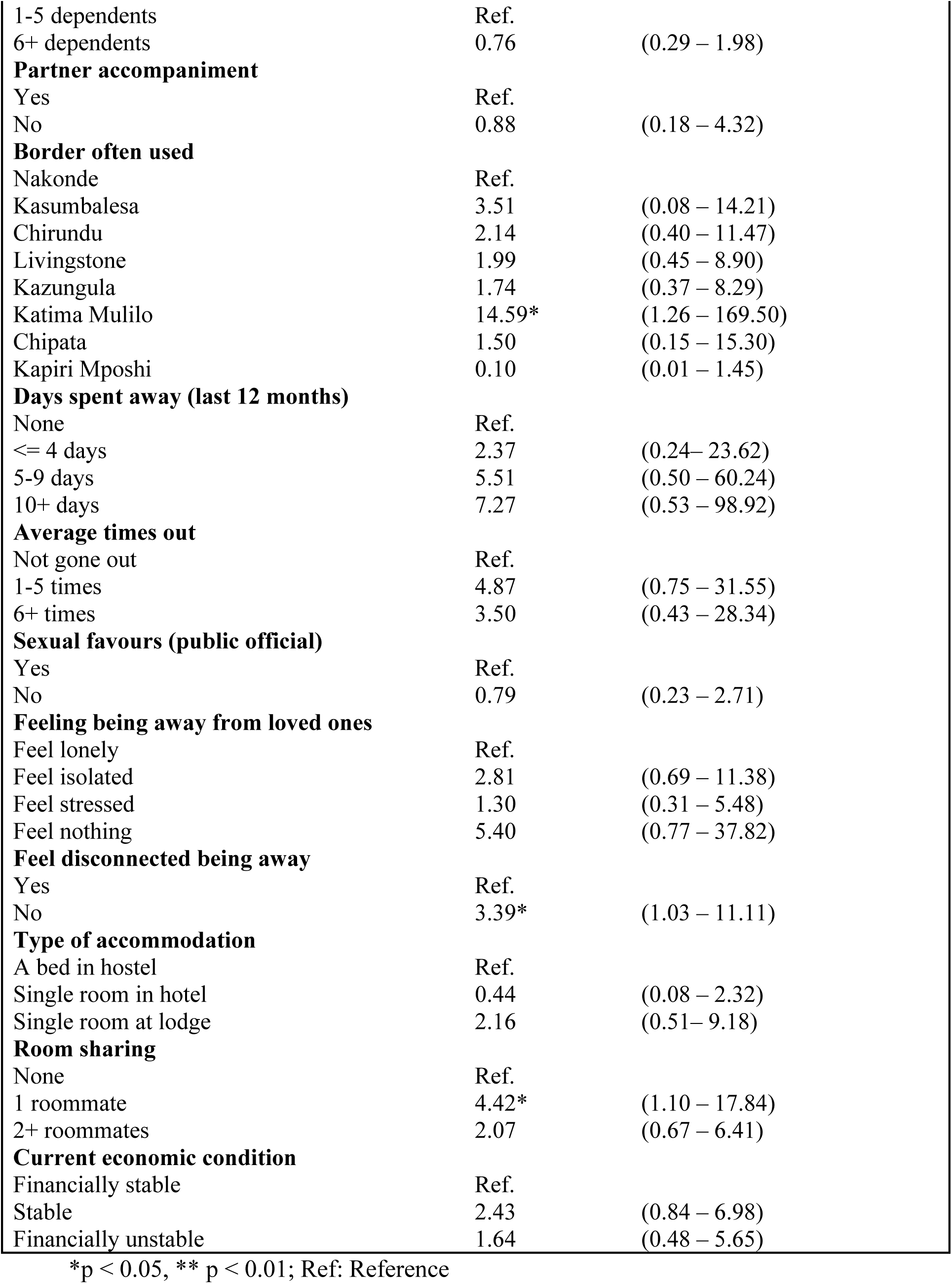
Adjusted binary logistic regression model for risky sexual behaviour by selected pre-migratory, migratory and post-migratory covariates.

### Pre-migratory factors

After adjusting for other covariates, source of start-up capital and period as a cross-border trader remained significant predictors of risky sexual behaviour. Women who used personal savings as start-up capital were 73% less likely to be in the high-risk group compared to those who relied on nuclear family capital (aOR = 0.27, 95% CI: 0.10-0.73, p < 0.01). With respect to trading experience, women who had been trading for 5-9 years were 79% less likely to report high-risk behaviour compared to those with 5 years or less experience (aOR = 0.21, 95% CI: 0.29-0.96, p < 0.05). The odds were also lower for those with 10+ years of experience (aOR = 0.16, 95% CI: 0.02-1.40), though this did not reach statistical significance. Initiator of trade, year of debut, age at debut, number of dependents, and partner accompaniment were not significantly associated with risky sexual behaviour in the adjusted model.

### Migratory factors

Among migratory variables, border crossing point was a significant predictor. Women who most often used the Katima Mulilo border were approximately 15 times more likely to be classified as high-risk compared to those using Nakonde (aOR = 14.59, 95% CI: 1.26-169.50, p < 0.05). The odds were also elevated for Kasumbalesa, Chirundu, Livingstone and Kazungula users, but these were not statistically significant. Days spent away in the last 12 months and average number of times out were not significantly associated with risk after adjustment, though the odds of high-risk behaviour increased with more days away and more trips. Similarly, being asked for sexual favours by public officials was not a significant predictor.

### Post-migratory factors

Two post-migratory factors were significantly associated with risky sexual behaviour. Women who reported “not” feeling disconnected while away from home had over three times higher odds of high-risk behaviour compared to those who felt disconnected (aOR = 3.39, 95% CI: 1.03-11.11, p < 0.05). Room sharing was also significant. Women who shared a room with one roommate had 4.4 times higher odds of being in the high-risk group compared to those who had no roommate (aOR = 4.42, 95% CI: 1.10-17.84, p < 0.05). The odds were also elevated for those sharing with 2+ roommates (aOR = 2.07, 95% CI: 0.67-6.41), but not statistically significant. Type of accommodation, feelings while being away, and current economic condition were not significantly associated with risky sexual behaviour in the adjusted model.

In summary, multivariate results indicated that financial independence through personal savings, longer trading experience, use of the Katima Mulilo border, not feeling disconnected while away, and sharing accommodation with one roommate were key factors independently associated with risky sexual behaviour among women informal cross-border traders.

## Discussion

The findings of this study show that risky sexual behaviour among women informal cross-border traders at the COMESA Market is influenced by factors across the pre-migratory, migratory, and post-migratory stages, consistent with migration and vulnerability theory [1]. This supports the view that vulnerability is not inherent to migrants, but is produced through interactions between individual characteristics and structural contexts encountered during mobility [8].

Pre-migratory factors such as economic dependence and source of start-up capital were significantly associated with risky sexual behaviour. Women who relied on family, friends, or informal lenders for capital were more likely to report transactional sex and inconsistent condom use. This aligns with evidence from the COMESA region showing that limited financial autonomy increases women’s reliance on sexual partnerships as a coping strategy during trade [2,3]. Similarly, the African Development Bank [4] and World Bank [5] note that economic pressure is a key driver of vulnerability among women traders in Africa, as it reduces bargaining power in sexual relationships. This suggests that interventions targeting financial inclusion and access to affordable credit may reduce sexual risk among women informal cross-border traders.

Migratory factors including frequency of travel and border crossing points also emerged as important. Women who crossed borders more than twice a month and those using busier, less regulated borders reported higher rates of risky sexual behaviour. This is consistent with literature showing that repeated mobility and transit through high-volume border posts increase exposure to informal economies and new sexual networks [6,9]. Migration studies in Southern Africa have similarly found that mobility disrupts social controls and increases contact with transient sexual partners, thereby elevating HIV risk [1,11]. The Southern African Development Community (SADC) regional strategy further highlights border areas as hotspots requiring targeted HIV prevention for mobile populations [13].

Post-migratory factors such as accommodation type and social isolation were also associated with risk. Women who stayed in shared lodges or hostels and those who reported feeling isolated at destination were more likely to engage in risky sexual behaviour. This mirrors findings by WIEGO [7] and USAID [10] that poor living conditions and weak social support at trading destinations increase vulnerability for women informal cross-border traders. Shared accommodation often lacks privacy and exposes women to peer norms that may normalise multiple partnerships or transactional sex. The IOM [12] also emphasises that destination environments shape health behaviours among mobile populations, particularly when access to health services is limited.

The overall prevalence of risky sexual behaviour observed in this study reflects broader national trends. The 2021 Zambia DHS reports that mobility and economic insecurity remain key drivers of HIV vulnerability among women [14]. At a regional level, UNAIDS [15] continues to identify mobile and key populations, including traders, as priority groups for HIV prevention.

Multivariate findings in this study align with migration and vulnerability theory, which posits that mobility exposes individuals to new social, economic, and physical environments that can heighten health risks, particularly when protective social ties are weakened. The increased odds of risky sexual behaviour among women who did not feel disconnected while away and those sharing accommodation with one roommate suggest that reduced social control and increased exposure to peer networks at destination may facilitate risk-taking, consistent with theory on social disorganisation during migration. Similarly, the substantially higher odds among users of the Katima Mulilo border point to how specific transit routes with distinct regulatory and social contexts can create unique vulnerability pathways. Conversely, the protective effects of using personal savings and having 5-9 years of trading experience support the theory’s emphasis on economic autonomy and migration maturity as buffers: financial independence reduces reliance on transactional relationships, while experience may foster coping strategies that mitigate exposure to risk. Collectively, these results underscore that vulnerability among women informal cross-border traders is not uniform, but shaped by the interplay of pre-migratory resources, migratory contexts, and post-migratory social environments.

Taken together, these findings underscore that risk among women informal cross-border traders cannot be addressed by individual-level behaviour change alone. As migration and vulnerability theory suggests, effective interventions must target all three stages: improving economic autonomy before travel, ensuring safer transit and regulation at borders, and providing health services and safe accommodation at destinations [1,19]. Programmes that integrate financial literacy, mobile health services, and peer support networks for women traders may be particularly effective in reducing vulnerability [3,7].

## Study limitations and Strengths

This study had several limitations that should be considered when interpreting the findings. First, the cross-sectional design limited the ability to establish causal relationships between migration-related factors and risky sexual behaviour. While associations could be identified, it was not possible to determine whether pre-migratory, migratory, or post-migratory conditions preceded the reported sexual risk behaviours. Second, data were collected through self-report, which may have been subjected to social desirability and recall bias, particularly given the sensitive nature of questions on sexual behaviour and transactional sex. Women may have underreported risky practices due to stigma. Third, the study was conducted among women informal cross-border traders at a single site, COMESA Market in Lusaka, which limits the generalisability of findings to women traders operating in other border markets or countries within the COMESA region. Despite these limitations, this study had many strengths. The use of migration and vulnerability theory provides a useful framework for understanding how mobility contexts shape risk, and the findings offer important insights for programming targeting mobile women traders. This study also uses a probability sample with a 99.8% response rate from a well-defined population of women traders at a major regional market. The use of a composite risk index and a migration-stage framework allowed us to examine how factors at departure, transit, and destination independently relate to sexual risk. In addition, the focus on informal cross-border traders addresses a gap in literature on mobile populations in Southern Africa who are often excluded from HIV programming.

## Conclusion

This study provides evidence that risky sexual behaviour is highly prevalent among women informal cross-border traders at the COMESA Market in Lusaka, with nearly two-thirds classified as high-risk. The findings demonstrate that vulnerability is not static, but shaped by factors across the migration process. Pre-migratory economic dependence, specific migratory routes, and post-migratory social and living conditions were all independently associated with sexual risk. Women who relied on personal savings and had moderate trading experience were less likely to engage in high-risk behaviour, highlighting the protective role of financial autonomy and migration maturity. Conversely, use of the Katima Mulilo border, lack of feelings of disconnection while away, and sharing accommodation with one roommate significantly increased the odds of high-risk behaviour. These results support migration and vulnerability theory, which posits that mobility creates new exposures when economic resources are limited and social controls are weakened. The study underscores that addressing sexual health among this population requires interventions that go beyond individual behaviour change to target the structural and contextual drivers of risk.We therefore recommend: 1) Promoting financial independence through savings groups and micro-credit; 2) Strengthening HIV prevention, protection, and health services at high-risk borders such as Katima Mulilo; 3) Improving access to safe, private accommodation and establishing peer-led health programmes in trader lodges; and 4) Conducting further longitudinal and multi-site studies are needed to better understand the temporal dynamics of risk and to validate these findings across different trading contexts and inform targeted interventions. Addressing structural and contextual drivers is critical to reducing HIV vulnerability among women traders.

## Declarations

### Consent for publication

Not applicable

### Availability of data and materials

The data are available upon request from the authors who is a PhD student at University of Zambia.

## Author Contributions

The conceptualisation of the project involved Simson Mwale, Musonda Lemba, and Million Phiri. Data curation was conducted by Simson Mwale and Musonda Lemba, with Simson Mwale also performing the formal analysis. Simson Mwale and Musonda Lemba developed the methodology. Simson Mwale was responsible for writing the original draft, and the review and editing of the work were undertaken by all the authors.

## Data Availability

The data are available upon request from the authors who is a PhD student at University of Zambia.

## Acknowledgements

The authors are grateful to the managers of COMESA market and women CBTA members who participated in this study.

